# Small apolipoprotein(a) isoforms may predict primary patency following peripheral arterial revascularization

**DOI:** 10.1101/2024.03.18.24304485

**Authors:** Marianna Pavlyha, Madeleine Hunter, Roman Nowygrod, Virenda Patel, Nicholas Morrissey, Danielle Bajakian, Yihao Li, Gissette Reyes-Soffer

## Abstract

**Background:** High lipoprotein (a) [Lp(a)] is associated with adverse limb events in patients undergoing lower extremity revascularization. Lp(a) levels are genetically pre-determined, with *LPA* gene encoding for two apolipoprotein (a) [apo(a)] isoforms. Isoform size variations are driven by the number of kringle IV type 2 (KIV-2) repeats. Lp(a) levels are inversely correlated with isoform size. In this study, we examined the role of Lp(a) levels, apo(a) size and inflammatory markers with lower extremity revascularization outcomes.

**Methods:** 25 subjects with chronic peripheral arterial disease (PAD), underwent open or endovascular lower extremity revascularization (mean age of 66.7±9.7 years; F=12, M=13; Black=8, Hispanic=5, and White=12). Pre- and post-operative medical history, self-reported symptoms, ankle brachial indices (ABIs), and lower extremity duplex ultrasounds were obtained. Plasma Lp(a), apoB100, lipid panel, and pro-inflammatory markers (IL-6, IL-18, hs-CRP, TNFα) were assayed preoperatively. Isoform size was estimated using gel electrophoresis and weighted isoform size (*wIS*) calculated based on % isoform expression. Firth logistic regression was used to examine the relationship between Lp(a) levels, and *wIS* with procedural outcomes: symptoms (better/worse), primary patency at 2-4 weeks, ABIs, and re-intervention within 3-6 months. We controlled for age, sex, history of diabetes, smoking, statin, antiplatelet and anticoagulation use.

**Results:** Median plasma Lp(a) level was 108 (44, 301) nmol/L. The mean apoB100 level was 168.0 ± 65.8 mg/dL. These values were not statistically different among races. We found no association between Lp(a) levels and w*IS* with measured plasma pro-inflammatory markers. However, smaller apo(a) *wIS* was associated with occlusion of the treated lesion(s) in the postoperative period [OR=1.97 (95% CI 1.01 - 3.86, p<0.05)]. The relationship of smaller apo(a) *wIS* with re-intervention was not as strong [OR=1.57 (95% CI 0.96 - 2.56), p=0.07]. We observed no association between *wIS* with patient reported symptoms or change in ABIs.

**Conclusions:** In this small study, subjects with smaller apo(a) isoform size undergoing peripheral arterial revascularization were more likely to experience occlusion in the perioperative period and/or require re-intervention. Larger cohort studies identifying the mechanism and validating these preliminary data are needed to improve understanding of long-term peripheral vascular outcomes.

**Key Findings:** 25 subjects with symptomatic PAD underwent open or endovascular lower extremity revascularization in a small cohort. Smaller apo(a) isoforms were associated with occlusion of the treated lesion(s) within 2-4 weeks [OR=1.97 (95% CI 1.01 - 3.86, p<0.05)], suggesting apo(a) isoform size as a predictor of primary patency in the early period after lower extremity intervention.

**Take Home Message:** Subjects with high Lp(a) levels, generally have smaller apo(a) isoform sizes. We find that, in this small cohort, patients undergoing peripheral arterial revascularization subjects with small isoforms are at an increased risk of treated vessel occlusion in the perioperative period.

**Table of Contents Summary:** Subjects with symptomatic PAD requiring lower extremity revascularization have high median Lp(a) levels. Individuals with smaller apo(a) weighted isoform size *(wIS)* have lower primary patency rates and/or require re-intervention.

## Introduction

Lipoprotein (a) [Lp(a)] is causal for the development of atherosclerotic cardiovascular disease (ASCVD) and atherothrombosis as shown in epidemiological, Mendelian Randomization, and genome wide association studies (GWAS) ^1^. Elevated Lp(a) levels are independently associated with incidence of major adverse cardiovascular events (MACE) and major adverse limb events (MALE) after lower extremity endovascular ^2^ or open surgery ^3^ while controlling for other risk factors. Despite this, measurement of Lp(a) levels is not standard of care in those undergoing vascular intervention. Lp(a) particles contain two main apolipoproteins: B100 [apoB100] and apolipoprotein(a) [apo(a)]. Individuals can express one or two apo(a) isoforms and these vary in size depending on the number of Kringle-IV type 2 (KIV-2) repeats that are encoded by the *LPA* gene. Plasma Lp(a) levels and apo(a) isoform size are inversely correlated, with smaller isoforms generally dominating^1^. There are well described mechanisms linking high levels of Lp(a) and atherosclerosis, including complement activation, inflammatory, and coagulation pathways^1^. Apo(a) can potentiate atherothrombosis through presence of lysine binding sites that allow accumulation of lipid in the arterial wall and activation of inflammatory pathways, owning to its binding of oxidized phospholipids (OxPL) ^4^ ^5^. High Lp(a) levels have also been association with enhanced expression of adhesion molecules in endothelial cells and pro-inflammatory interleukins in monocytes and macrophages ^6^. Finally, apo(a) has evolved from the plasminogen gene through duplication and remodeling ^7^ and its homology with plasminogen allows it to compete for its binding site, inhibiting thrombolysis and indirectly promoting a prothrombotic state. These mechanisms are of interest in vascular patency.

Numerous studies show that high Lp(a) levels are associated with increased incidence of claudication, symptoms progression, re-stenosis after intervention, hospitalization, and limb amputation ^8^, and those with high Lp(a) have a higher risk of combined PAD outcomes after adjusting for other traditional risk-factors. Few studies, however, have looked at the association between apo(a) isoform size and peripheral arterial surgical outcomes. Available data suggests that both increased Lp(a) levels and the presence of small apo(a) isoforms are associated with the increased incidence of symptomatic PAD ^9^ ^10^. Whereas small apo(a) isoforms were shown to be independently associated with three-fold increase in risk of major adverse cardiovascular events after coronary artery bypass grafting (CABG) at long term follow up ^11^, the role of this Lp(a) component on lower limb outcomes have not been explored. In view of previous findings and needs in the field, we developed a study using available stored plasma samples and outcomes data on subjects with PAD who underwent lower extremity revascularization, yet they had no history of Lp(a) measurement. We examined the relationships between apo(a) isoform size, primary patency, and rate of re-intervention. In addition, given apo(a)’s role in propagating the inflammatory cascade implicated in atherosclerotic plaque formation^12^ ^13^ ^14^, the relationship of pro-inflammatory markers (IL-6, IL-18, hs-CRP, TNFα) with apo(a) size and surgical outcomes were investigated.

There are current studies focused on lowering Lp(a) by targeting the apo(a) component in phase 3 of development ^15^, highlighting the need to understand which populations are at increased disease risk and would benefit from treatment. Our small study provides preliminary insights that can be used to design larger studies in patients with advanced PAD. These studies may assist with improving risk prediction, stratification, and medical optimization of surgical patients with high Lp(a).

## Methods

We analyzed data from all 27 participants recruited from the Columbia University Irving Medical Center (CUIMC) Vascular Surgery Clinic into a prospective cohort study [Anthropometric and Biomarker Assay, and Lower Extremity Arterial Revascularization (ABALEAR)] between June 2018 and March 2021, **Supplemental Figure 2**. The study protocol was approved by the CUIMC Institutional Review Board (AAAL7756). Inclusion criteria encompassed adult subjects with a clinical diagnosis of PAD, who were planned to undergo non-emergent revascularization (open or endovascular) of the affected lower extremity. Those with ischemic wounds, active infectious process, or cardiovascular intervention within 12 months of enrollment were excluded. Two of the participants who underwent only diagnostic angiogram were also excluded from final analysis. The type of intervention was at the discretion of the treating provider.

### Recruitment/Enrollment

Subjects that met inclusion and exclusion criteria were contacted prior to their scheduled operation and informed consent was obtained. Medical and social history were collected by a licensed health professional, including demographic information (age, sex, self-reported race-ethnicity (SRRE)), history of smoking, hypertension (HTN), diabetes, carotid artery disease (CAD) or dyslipidemia. Medication use [antihypertensives, statins, antiplatelet (AP), direct oral anticoagulant (DOAC)] and symptoms of rest pain or claudication (ambulatory distance in city blocks) were recorded. A pre-operative physical exam was performed, which included distal pulses, a lower extremity arterial duplex ultrasound (US), ankle brachial index (ABI), and body mass index (BMI). On the day of the surgical procedure, arterial blood samples were collected using EDTA tubes; plasma was separated and stored at −80◦C.

### Study Follow-up (2-4 weeks and 3-6 months)

Participants were seen in the office for their post-operative visits by their respective providers. During these visits, physical exam (pulses, US, ABIs) to assess for primary patency and the trajectory of self-reported subjective symptoms were determined based on presence or resolution of rest pain and changes in walking distance. These were then categorized as ‘improved,’ ‘worsened’ or ‘stayed the same.’ In cases of multiple vessels intervention, absent blood flow in the most proximal intervened vessel constituted occlusion. If multiple vessels below the popliteal artery trifurcation were intervened on, multiphasic in-line flow preserved in at least one of the vessels constituted patency. Palpable pulses over the graft and distally constituted patency. Two subjects did not have a complete US assessment at first follow-up and were excluded from sub-analysis. Change in ABI^16^ measurements were categorized as improved from pre-operative measurement vs not improved (worsened or the same). Of note, ABI measurements falsely elevated due to calcified non-compressible vessels, incomplete, or missing for at least one of the visits were excluded from the sub-analysis. Hence, the available ABI data is limited. Toe pressures were not available on this cohort. In addition to the above, the 3-6 months visit included records of any interval interventions on the ipsilateral extremity.

Plasma Lp(a), apoB100, lipids [triglycerides (TG), total cholesterol(C), high density lipoprotein (HDL)] were automatically measured on Integra 400 plus (Roche). Plasma LDL-C levels were calculated using the Friedewald formula (one subject had a TG level >400mg/dL and was not reported). Plasma apoB100 levels were measured by a human enzyme-linked immunosorbent assay (ELISA) from Mabtech, Inc, Cincinnati, OH. Lp(a) plasma concentration were measured using the isoform-independent sandwich ELISA developed by the Northwest Lipid Metabolism and Diabetes Research Laboratory at Medpace, Inc. Apo(a) isoform size measurements were performed by the same laboratory using agarose gel electrophoresis. ChemiDoc MP Imaging System was used to determine the isoforms using in-house standards and relative expression of each isoform was reported. Weighted isoform size (*wIS*) was calculated using the % isoform expression (two isoform masses were multiplied by their relative expression). Pro-inflammatory markers Interleukin-6 (IL6), Interleukin-18 (IL-18) were measured using ELISA kits (D6050, R&D Systems, Minneapolis, MN) and (DY318-05, R&D Systems, Minneapolis, MN), respectively. High sensitivity C-reactive protein (hs-CRP) and Tumor Necrosis Factor alpha (TNFα) were measured on an Integra400plus (Roche) using standardized methods at the Irving Institute for Clinical and Translational Research Biomarkers laboratory at CUIMC.

### Statistical Analysis

All data were analyzed using standard R software (Version 4.2.2). Normally distributed variables were summarized by means and standard deviations (SD). Lp(a) levels and TGs were summarized by median and interquartile range. The relationship of Lp(a) levels with *wIS* in the three SRRE groups was studied by Analysis of Variance (ANOVA). Racial differences in Lp(a) levels and *wIS* were assessed using the median test. Due to small samples size, an implementation based on the Firth logistic regression ^17^was chosen to examine the relationship between *wIS* with procedural outcomes: change in symptoms, primary patency of treated lesion(s), change in ABIs, and re-intervention within 3-6 months. Results were reported as odds ratios (OR). Sex, age, smoking status, history of diabetes, statin, antiplatelet and DOAC use were considered as potential confounders, according to the literature, and were included in the regression ^18^ ^19^ ^20^ ^21^. HTN and BMI were not integrated into the model based on their role in published studies of surgical outcomes ^22^ ^23^. We did not control for history of CAD, as literature shows that most individuals with diagnosis of PAD have some degree of CAD and at least 60% have significant CAD^24^..

## Results

### Baseline Characteristics

Subject demographics, lipid and lipoprotein levels are shown in **Table I**. The study population included 12 females and 13 males with a mean age of 66.7±9.7 years. Our cohort was multiethnic with 8 Blacks, 5 Hispanics, and 12 Whites, representative of the available population at our institution. All subjects were Rutherford stage 3 and 4. 5 subjects underwent open revascularization and 20, endovascular. Majority of the subjects underwent infra-inguinal intervention (N=23) and 6 required re-intervention within 3-6 months. Individuals were mildly overweight and lipid levels consistent with a population being treated for dyslipidemia. Despite use of lipid lowering therapies, the mean apoB100 levels were 168.0 ± 65.8 mg/dL. Median plasma Lp(a) level was 108 (44, 301) nmol/L. Primary patency rates at 2-4 weeks and 3-6 months were 76% and 68%. Rate of re-intervention measured at 3–6-months visit was 24%.

**Table I.**
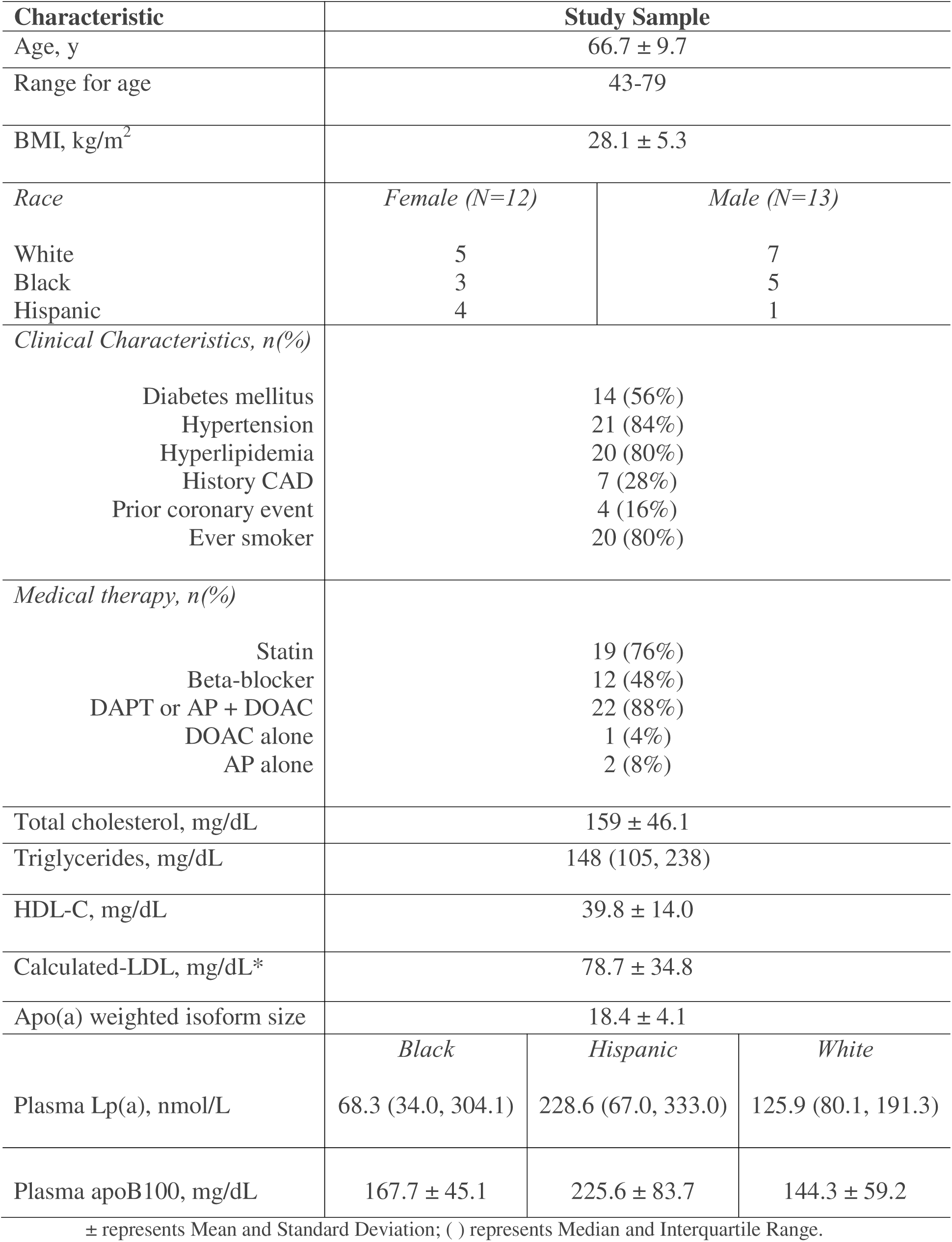
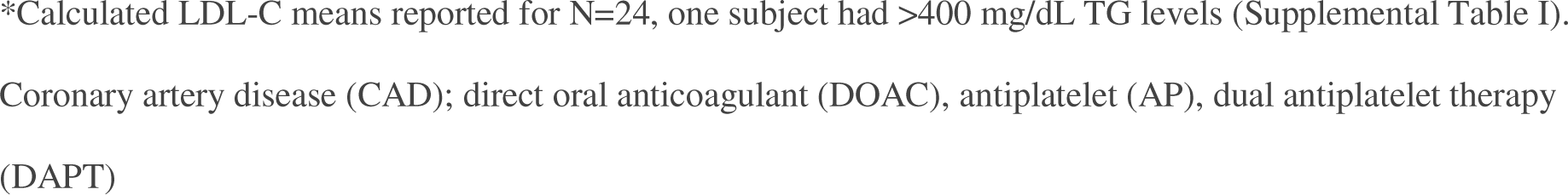
Baseline characteristics, lipid, and lipoprotein levels.

| Characteristic | Study Sample |  |  |
| --- | --- | --- | --- |
| Age, y | 66.7 ± 9.7 |  |  |
| Range for age | 43-79 |  |  |
| BMI, kg/m<sup>2</sup> | 28.1 ± 5.3 |  |  |
| Race | Female (N=12) | Male (N=13) |  |
| White | 5 | 7 |  |
| Black | 3 | 5 |  |
| Hispanic | 4 | 1 |  |
| Clinical Characteristics, n(%) |  |  |  |
| Diabetes mellitus | 14 (56%) |  |  |
| Hypertension | 21 (84%) |  |  |
| Hyperlipidemia | 20 (80%) |  |  |
| History CAD | 7 (28%) |  |  |
| Prior coronary event | 4 (16%) |  |  |
| Ever smoker | 20 (80%) |  |  |
| Medical therapy, n(%) |  |  |  |
| Statin | 19 (76%) |  |  |
| Beta-blocker | 12 (48%) |  |  |
| DAPT or AP + DOAC | 22 (88%) |  |  |
| DOAC alone | 1 (4%) |  |  |
| AP alone | 2 (8%) |  |  |
| Total cholesterol, mg/dL | 159 ± 46.1 |  |  |
| Triglycerides, mg/dL | 148 (105, 238) |  |  |
| HDL-C, mg/dL | 39.8 ± 14.0 |  |  |
| Calculated-LDL, mg/dL* | 78.7 ± 34.8 |  |  |
| Apo(a) weighted isoform size | 18.4 ± 4.1 |  |  |
|  | Black | Hispanic | White |
| Plasma Lp(a), nmol/L | 68.3 (34.0, 304.1) | 228.6 (67.0, 333.0) | 125.9 (80.1, 191.3) |
| Plasma apoB100, mg/dL | 167.7 ± 45.1 | 225.6 ± 83.7 | 144.3 ± 59.2 |

\*Calculated LDL-C means reported for N=24, one subject had >400 mg/dL TG levels (Supplemental Table I).
Coronary artery disease (CAD); direct oral anticoagulant (DOAC), antiplatelet (AP), dual antiplatelet therapy (DAPT)

### Lp(a) levels and Isoform Size

Previous reports show that high Lp(a) levels are associated with worse outcomes after surgical intervention. We found that most of the individuals in our cohort had high Lp(a) levels. The mean *wIS* was 18.4 ± 4.1 and as expected, there was an inverse relationship between Lp(a) levels and *wIS* (R^2^ = 0.20; p=0.02), **Figure 1**. Most individuals express two apo(a) isoforms, however in our small diverse cohort, we found that half (N=12) individuals expressed only a single isoform, **Supplemental Table I**, with predominance of the small isoform, **Figure 2**.

**Figure 1.**
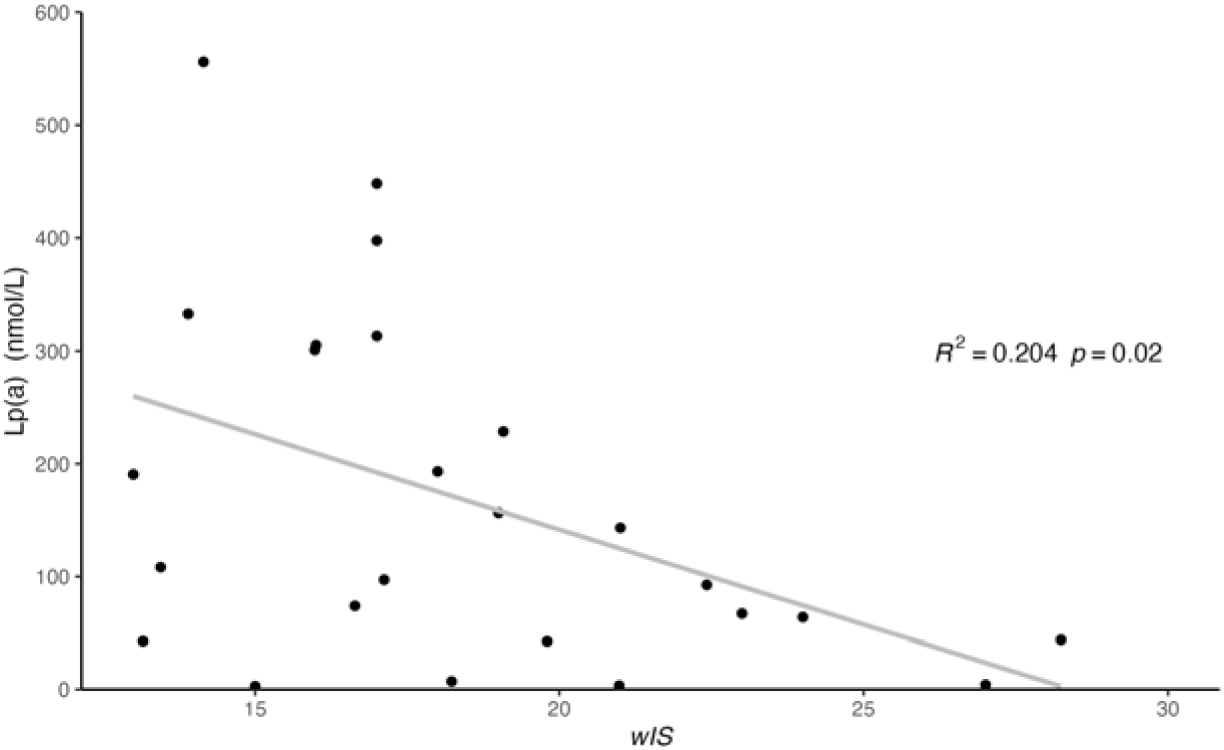
Negative relationship between Lp(a) levels and *wIS*. Lp(a): Lipoprotein(a); *wIS*: weighted Isoform Size.

**Figure 2.**
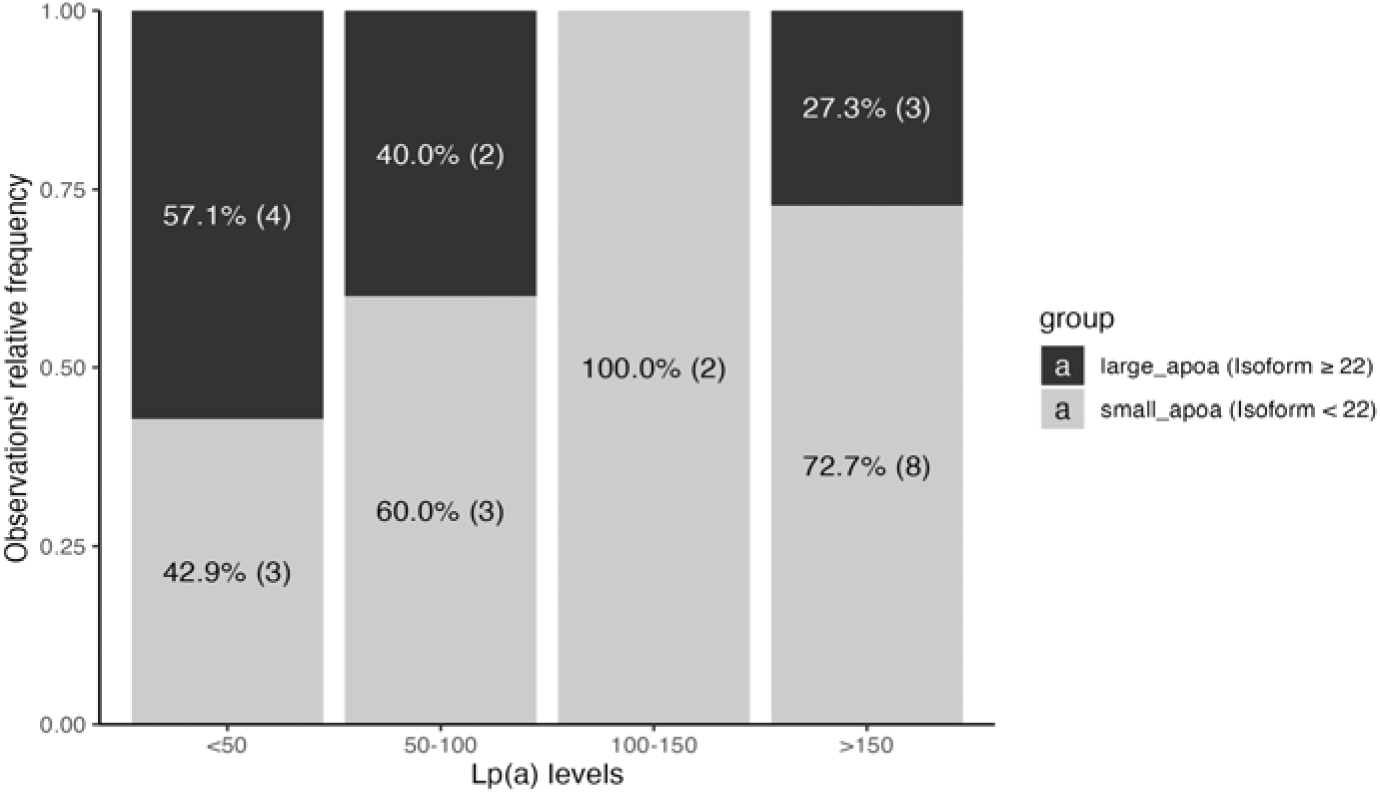
Apo(a) size distribution by Lp(a) level quartiles (nmol/L) Lp(a): Lipoprotein(a). Gray: small apo(a) isoforms; Black: large apo(a) isoforms. All data presented as percentages of study subjects and absolute numbers in ().

### Relationship of Lp(a) levels and Isoforms with surgical outcomes

We did not find significant associations between Lp(a) levels and post-surgical symptoms, ABI, primary patency, or re-intervention, **Table IIb**. Surgery type did not affect this result. However, *wIS* was significantly correlated with primary patency at 2-4 weeks [OR=1.97 (95% CI 1.01 - 3.86, p<0.05)], **Table IIa**. Controlling for SRRE did not change these study outcomes. We observed a trend between *wIS* and re-intervention in the 3-6 months period [OR=1.57 (95% CI 0.96 - 2.56), p=0.07)]. When we used predominant apo(a) isoform size as the primary variable in our model, and adjusted for plasma Lp(a) levels, our observed effect of apo(a) size on primary patency remained statistically significant [OR=2.01, (CI 1.03 - 3.93) p<0.05], suggesting that apo(a) was a stronger a predictor of outcomes compared to Lp(a) levels in this cohort.

**Table IIa.** Effects of apo(a) *wIS* on surgical outcomes.

| Outcome | Sample (N) | Firth Model Estimate (95% CI) | p Value |
| --- | --- | --- | --- |
| Δ Symptoms | 25 | 0.25 (-1.01, 1.50) | p=0.70 |
| Primary patency | 23 | **-2.81 (-5.57, -0.04)** | **p <0.05\*** |
| Δ ABI | 14 | 0.30 (-2.43, 3.01) | p=0.84 |
| Re-Intervention | 25 | **-0.53 (-2.59, 1.54)** | **p=0.07** |

**Table IIb.** Effects of Lp(a) on surgical outcomes.

| Outcome | Sample (N) | Firth Model Estimate (95% CI) | p Value |
| --- | --- | --- | --- |
| Δ Symptoms | 25 | 0.17 (-0.89, 1.23) | p=0.75 |
| Primary patency | 23 | 0.32 (-0.68, 1.33) | p=0.53 |
| Δ ABI | 14 | 0.92 (-1.57, 3.41) | p=0.47 |
| Re-intervention | 25 | 0.13 (-0.89, 1.15) | p=0.80 |

The type of surgical procedure could have affected our study outcomes, however when we assessed the relationship between *wIS* isoform size and primary patency for subjects that only underwent endovascular intervention, we noted a similar trend [OR=1.67 (95% CI 0.95 - 2.94), p=0.08]. We did not see an association between *wIS* and patient reported symptoms or change in ABIs at 3-6 months. It is notable, however, that our sample size for analysis of ABIs was much smaller (N=14) due to inability of all subjects to undergo the test reliably (ex. pain, non-compliant calcified vessels resulting in falsely elevated results) or missing measurement from some visits.

As expected, inflammatory markers IL-18 and hs-CRP were higher in this cohort when compared to published levels in healthy cohorts, **Table III**. However, we found no significant relationship between plasma Lp(a) levels or *wIS* with IL-6, IL-18, hs-CRP, or TNFα. Additionally, inflammatory markers were not associated with our surgical outcomes.

**Table III.** Comparison of inflammatory marker levels in Study Subjects and Healthy Controls.

| **Marker** | **PAD Cohort** | **\*Healthy Controls** |
| --- | --- | --- |
| IL-6 (pg/mL) | 3.3±2.5 | 0.01-11.5 <sup>38</sup> |
| IL-18 (pg/mL) | 274.2±147.5 | 80–120 <sup>39</sup> |
| hs-CRP (mg/dL) | 3.6±4.6 | <0.3 <sup>40</sup> |
| TNFα (pg/mL) | 1.4±0.7 | 0 - 2.2 |

Factor. Study Cohort N=25. \*Healthy controls from published literature

## Discussion

High Lp(a) levels are associated with adverse limb events in patients with symptomatic PAD undergoing lower extremity revascularization ^25^ ^26^ ^2^. International guidelines (Canadian ^27^ and ECC ^28^) advocate for all individuals to have Lp(a) measured once in a lifetime. In the United States, the American Heart Association (AHA) suggest using Lp(a) levels as a ‘risk enhancer’^29^. However, there are no specific guidelines for Lp(a) measurements in patients with advanced PAD that are referred for surgical intervention. All subjects in our study have been diagnosed with at least one metabolic disease, however, they were not screened for Lp(a) by their primary providers; and the majority were found to have elevated levels.

Although our study sample is small, which limits the generalizability of the results, it features a good representation based on gender and race. These data can be used to develop well controlled cross-sectional studies that can examine the role of apo(a) in larger populations with PAD. Most individuals at risk or that have ASCVD, obtain laboratory measurement of low-density lipoprotein cholesterol (LDL-C), an apoB100 containing particle. Physicians rely on these measurements for prescribing lipid therapies, such as statins, in clinical practice. We observed that in our cohort, 76% of individuals were prescribed statins. While statins significantly lower LDL-C, they tend to increase Lp(a) by about 11% ^1^. It is important to note, however, that the laboratory levels of LDL-C in plasma are known to be confounded by the presence of Lp(a)-C^30^, which contains both apoB100 and apo(a). The reason for this overlap lies in similar plasma densities between Lp(a) and LDL particles. Thus, in patients with high levels of Lp(a) the actual LDL-C is lower from the reported value and some investigators believe apoB100 to be a more accurate measure of cardiovascular risk compared to LDL-C^31^. This is important, because most vascular patients are prescribed statins, which works via increased LDL receptor expression, facilitating clearance of LDL-C, but not Lp(a). Patients that have elevated LDL-C while on optimal LDL lowering therapy would benefit from having their Lp(a) measured and treated, as there is a high probability that the increased cholesterol is carried by Lp(a) particles. We observe this in some patients in our population as well and this may explain why some patients, despite LDL-C lowering by statins, remain at risk^32^. Two out of 19 subjects in the study who were taking statins had above normal LDL-C levels; both had Lp(a) measurements > 200 nmol/L. In this population where Lp(a) is high, this particle may carry most of the cholesterol.

Whereas there has been great interest in Lp(a) levels and surgical outcomes, few studies have looked at the role of apo(a) size in vascular surgery, and specifically, there are no existing reports on patency after lower extremity revascularization. The *LPA* gene encodes for two apo(a) isoforms that comprise the Lp(a) particle. Variations in size of the apo(a) isoforms are driven by the number of KIV-2 repeats. Small number of KIV-2 repeats lead to small apo(a) size and are associated with higher Lp(a) levels, posing an increased risk for cardiovascular disease, including PAD ^9^. Similar to other cohorts^1^, the current study finds an inverse association between plasma Lp(a) levels and apo(a) allele size, with smaller isoforms associated with higher Lp(a) levels^33^. Whereas ∼20% of the general population is expected to have high Lp(a) and express two apo(a) isoforms^1^, close to half of our subjects with high median Lp(a) expressed only one isoform size, **Supplemental Table I**. Previous studies in individuals with PAD showed that both increased Lp(a) serum concentrations (75th percentile) and the small apo(a) phenotype are strongly associated with symptomatic PAD, independent of other confounders ^9^. We did not see any significant correlation between Lp(a) levels and outcomes, but when controlled for Lp(a) levels, the predictive association of *wIS* with primary patency was still statistically significant. Although data in the literature regarding disease predictability of Lp(a) vs apo(a) is conflicting, there is evidence that the strength of the association is higher for apo(a) isoform size in certain diseases^34^. To our knowledge, there are no reports assessing the correlation between apo(a) size and primary patency. Our preliminary study finds that small apo(a) size strongly correlates with primary patency within 2-4 weeks with a trend towards a higher rate or re-intervention at 3-6 months. Although more comprehensive evaluation of the prognostic relationship between apo(a) size and these outcomes of peripheral arterial interventions would be served in a randomized or cross-sectional study, these results support what is already known about apo(a)’s mode of action. We postulate that the probable mechanisms lie in the apo(a)’s kringle structure having homology with fibrinolytic proenzyme plasminogen, but inactive in fibrinolysis. Small apo(a) isoform size resulting from low *LPA* KIV-2 number of repeats can slow fibrinolysis and thus indirectly promote thrombosis, leading to increased risk of perioperative complications such as graft occlusion or thromboembolic events ^35^. Additionally, apo(a) potentiates thrombosis by binding and inactivating tissue factor pathway inhibitor (TFPI)^36^. This is especially important, as many of peripheral arterial intervention are performed to aid in wound healing and this period may be imperative for patients with tissue loss. These pro-thrombotic action of the apo(a) could also explain the high rate of early re-intervention in this cohort of patients with high Lp(a), particularly as 5 our 6 subjects in the group requiring re-intervention had small apo(a), **Supplemental Figure 1.**

A study from a UK biobank studied approximately 460Lthousand participants and showed significant differences in Lp(a) concentrations across racial subgroups, specifically with higher Lp(a) levels seen within South Asian and Black population compared to Whites ^37^. Although on average Hispanics had higher values than Whites in our cohort, there we no statistically significant differences in Lp(a) values and apo(a) size among different races and race had no impact on most surgical outcomes. This could be due to our limited sample size.

Some of Lp(a)’s role in disease risk has been linked to inflammatory profiles. Specifically, elevated TNF-α, IL-6, and CRP levels^12,38^ at baseline correlate with worse vascular outcomes in patients with diabetes, PAD, and chronic limb-threatening ischemia undergoing endovascular procedures ^14^. We did not find any significant correlations with circulating inflammatory markers and Lp(a), *wIS* or surgical outcomes, however, we did not measure OxPLs which may add new information in PAD patients as it has in other described populations ^39^. Of note, it has also been reported that statins decreased the serum levels of hs-CRP^12^ and downregulate IL-6^13^. Majority of our subject were taking high-dose statins (which also tend to increase Lp(a)) and may be contributing to our observations.

Although there are no currently available targeted therapies to treat high Lp(a), some agents available in the United States that lower apoB100 and LDL-C also decrease Lp(a) modestly, such as niacin, Ezetimibe, and PSCK9 inhibitors. The FDA has also recently approved lipoprotein apheresis for high-risk cases with Lp(a) >60mg/dL and LDL-C >100 mg/dL and with either documented CAD or PAD^1^. The two apo(a) targeted therapies, antisense oligonucleotides and siRNA and are also in phase 3 studies are expected to report their initial results in 2024. While we wait for this data, new studies now suggest the benefit of DAPT in patients with high Lp(a)^40^. Particularly, recent outcomes in these patients undergoing percutaneous coronary intervention (PCI), showed that prolonged DAPT (>1 year) reduced ischemic cardiovascular events, all-cause mortality, cardiac mortality, and definite/probable stent thrombosis, without increase in clinically relevant bleeding risk compared with ≤ 1-year DAPT ^41^. Data from this small study cohort raise the question whether we need to rethink risk assessment and post-operative management in patients high Lp(a) and small apo(a) size undergoing lower extremity interventions. As the importance of Lp(a) becomes more recognized in clinical practice^37,42^, and we review cardiovascular outcome data from phase 3 apo(a) targeted treatments, developing larger studies that have more stringent control of patient selection and type of intervention would serve as the next step in validating the generalizability of the current findings.

Understanding the impact of apo(a) isoform size on Lp(a) levels and surgical outcomes may guide risk stratification and therapeutic decision-making when it comes to pre-operative optimization and medical management post-operatively. Lp(a) levels are stable throughout lifetime. Although measuring apo(a) size may not be feasible in all patients and it currently not used in clinical practice, as genetic testing becomes more widely available, assessing KIV-2 size associated with the *LPA* gene may help identify individuals at higher risk of developing PAD and worse outcomes. Given Lp(a) genetic predisposition and low variability of levels throughout lifetime, one measurement may be sufficient and is the standard of care in many European countries^28^.

Further research in larger cohorts is needed to establish a more robust understanding of the mechanisms and impact of apo(a) isoform size on surgical outcomes in patients PAD. Randomized clinical trials to explore potential medical therapy strategies to mitigate associated risks would aid in providing guidelines for perioperative management of these individuals.

## Study Limitations

Overall, this was a sample of convenience. Subject samples and prospectively collected data from ongoing biomarker study (ABALEAR) were available to our lipid team to examine additional affecters on surgical outcomes. Although subject baseline characteristics are comparable to larger cohorts, we acknowledge that sample size is small and significantly limit the generalizability of the results. We used firth logistics regression to account for small sample size. Additionally, the cohort is heterogenous in terms of presenting symptoms with surgical interventions geared towards each patient’s needs, but ultimately ‘no leg is alike,’ and the cohort represents a diverse spectrum of lower extremity disease and intervention seen in vascular surgery clinics. Additionally, due to limited size, associations between peripheral interventions and Lp(a) levels/apo(a) size was done without correction for multiple testing.

## Conclusion

In this limited cohort, most subjects with chronic symptomatic PAD had elevated Lp(a) levels without being screened by their medical providers outside of the study, pointing to the need to increase awareness for Lp(a) screening in vascular patients. Small apo(a) isoform expression could be an important risk predictor not only for the development of PAD, which has already been established in literature, but also as a driver of worse short term surgical outcomes. Further studies identifying the mechanism and relationship of these findings to long-term peripheral vascular outcomes are warranted, specifically identifying the role of apo(a) size among different sexes and races in larger well controlled cohorts.

## Data Availability

All data produced in the present work are contained in the manuscript

## Acknowledgements

We would like to acknowledge Anastasiya Matveyenko^1^and Srinivas Cherukuri^1^for their assistance with study conduction and laboratory measurements. **Funding** for this study was provided by T32HL007343 (M.P.), UL1TR001873 (G.R.S), partial funding for MP stipend and manuscript processing funds were supported by a donation from Robin Chemers Neustein to the Reyes-Soffer Laboratory.

## Supplemental Data

**Supplemental Table I.** Individual Subject Data.

| Subject Number | Lp(a)<br>(nmol/L) | Isoform 1 | %<br>Isoform 1 | Isoform 2 | %<br>Isoform 2 | wIS | SRRE |
| --- | --- | --- | --- | --- | --- | --- | --- |
| 1 | 300.99 | 14 | 8% | 25 | 18% | 15.98 | B |
| 2 | 97.10 | 17 | 97% | 21 | 3% | 17.12 | W |
| 3 | 92.56 | 20 | 78% | 31 | 22% | 22.42 | W |
| 4 | 3.40 | 19 | 82% | 30 | 18% | 20.98 | B |
| 5 | 4.12 | 27 | 100% |  |  | 27 | B |
| 6 | 73.82 | 14 | 78% | 26 | 22% | 16.64 | B |
| 7 | 193.26 | 18 | 100% |  |  | 18 | W |
| 8 | 190.63 | 13 | 100% |  |  | 13 | W |
| 9 | 228.60 | 18 | 82% | 24 | 18% | 19.08 | H |
| 10 | 43.91 | 28 | 96% | 34 | 4% | 28.24 | B |
| 11 | 397.62 | 17 | 100% |  |  | 17 | H |
| 12* | 305.28 | 16 | 100% |  |  | 16 | W |
| 13 | 108.34 | 13 | 91% | 18 | 9% | 13.45 | W |
| 14 | 448.08 | 17 | 100% |  |  | 17 | W |
| 15 | 332.96 | 14 | 90% | 13 | 10% | 13.9 | H |
| 16 | 42.62 | 13 | 99% | 29 | 1% | 13.16 | W |
| 17 | 313.38 | 17 | 100% |  |  | 17 | B |
| 18 | <2.80 | 15 | 100% |  |  | 15 | W |
| 19 | 63.84 | 24 | 100% |  |  | 24 | B |
| 20 | 42.47 | 19 | 84% | 24 | 16% | 19 | H |
| 21 | 156.73 | 19 | 100% |  |  | 19 | W |
| 22 | 67.04 | 23 | 100% |  |  | 23 | H |
| 23 | 7.05 | 15 | 81% | 32 | 19% | 18.23 | W |
| 24 | 143.43 | 21 | 100% |  |  | 21 | W |
| 25 | 556.02 | 14 | 99% | 29 | 1% | 14.15 | B |

**Supplemental Figure 1.**
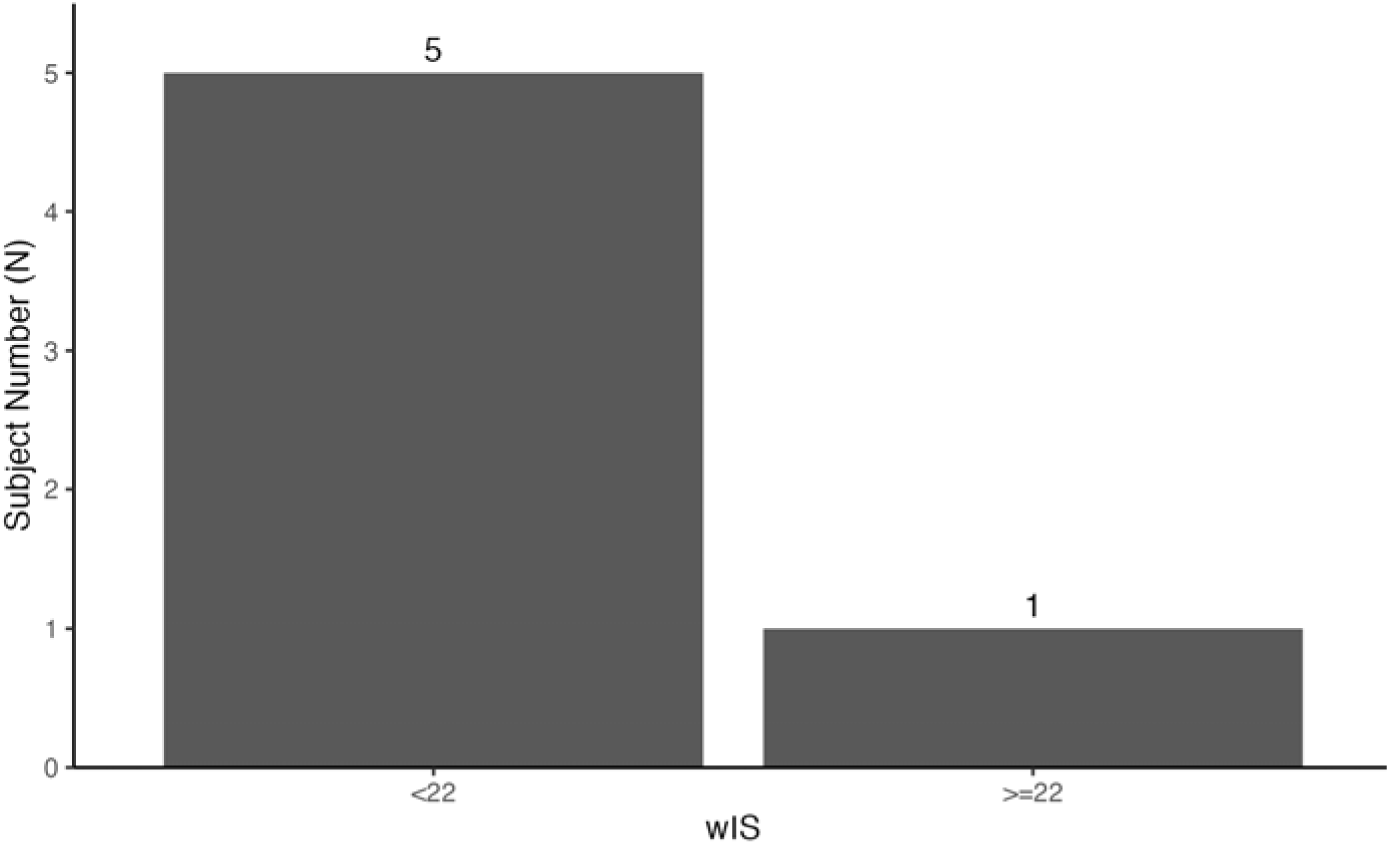
Apo(a) weighted isoform sizes in re-operative patients. wIS – weighted Isoform Size, Subject Number (N) underwent re-operation at 3-6 months.

**Supplemental Figure 2.**
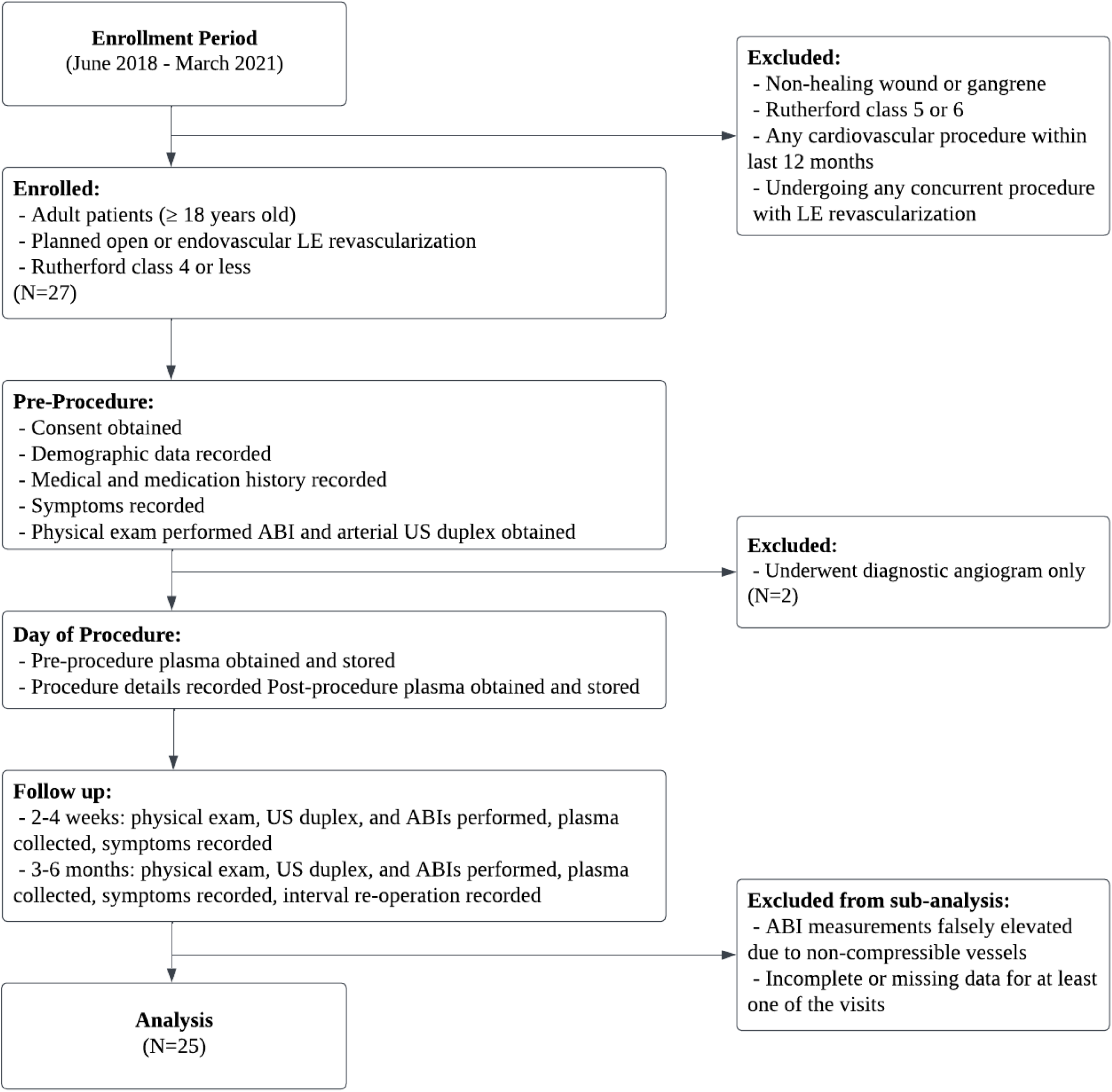
ABALEAR study CONSORT flow diagram. LE—lower extremity; US—ultrasound; ABI—ankle brachial index; N—sample number

